# Rapid review of government issued documents relevant to mitigation of COVID-19 in the US food manufacturing and processing industry

**DOI:** 10.1101/2022.02.25.22271516

**Authors:** Ziqian Chen, Ece Bulut, Aljoša Trmčić, Renata Ivanek

## Abstract

We surveyed publicly available records published by the United States (US) government between the start of the Coronavirus Disease 2019 (COVID-19) pandemic and September 30^th^, 2021, to identify documents containing resources or guidelines about COVID-19 mitigation relevant to the US food manufacturing and processing industry (hereafter referred to as “the food processing industry”). Among 36 documents identified and reviewed (including 35 from government agencies and one from a relevant professional association), we extracted 19 categories of mitigation strategies covering the themes of employee biosafety, surveillance, vaccination, social distancing, and worker education. We concluded that the priority of COVID-19 mitigation in the food processing industry was to protect the health and safety of industry workers while maintaining food supply chain resilience to minimize disturbance in the food market and avoid food crisis. A collated list of the identified documents and their comprehensive review will (i) aid researchers and public health workers in interpreting the potential impacts of the recommended mitigations on the epidemiology of the disease among workers in the food processing industry and (ii) help the food processing industry sort out the most essential strategies to take in face of a pandemic.

## INTRODUCTION

Since the first detected case in the United States (US) in early March 2020, the Coronavirus Disease 2019 (COVID-19) pandemic has significantly impacted many aspects of the society and many sectors of the US economy *(62)*. Specifically, the food manufacturing and processing industry (hereafter referred to as “the food processing industry” for short) has been heavily affected by labor shortages and supply issues *(44)*, which is of a high concern as food is a basic necessity of humans. One of the first documented large COVID-19 outbreaks in the food processing industry occurred at the Smithfield Foods pork processing plant in Sioux Falls, South Dakota, which resulted in 929 (26%) cases among its 3,635 employees *(38, 50)*. As of September 2021, more than 1,400 COVID-19 outbreaks at food processing and meatpacking facilities and more than 77,900 cases among workers in those facilities had been reported *(35)*. High transmission rate and a large number of cases in the processing facilities have forced many factories into temporary closure due to COVID-19 infections or measures to control the infection (e.g., quarantine) among workers, which has impacted countless employees and their families and threatened the normal functioning of the food supply chain *(37)*. The shortages of labor and closures of processing facilities have also negatively affected the livestock sector. Namely, the reduced capacity of the food processing sector resulted in a number of animal production, health, and welfare issues, such as overstocking of farms and the related stress and risk of animal diseases, and efforts to reduce the production including through drastic measures such as culling of animals and induced abortions *(40, 43)*. As the pandemic has placed significant stress on the national food supply chain, the food processing industry is recognized now more than ever as a critical infrastructure for preserving normal societal function.

To help combat the negative effects of the COVID-19 pandemic, the Centers for Disease Control and Prevention (CDC) rapidly responded with a series of documents to help guide mitigation efforts in the community, including in the food processing industry. On April 3^rd^, 2020, the CDC published ‘Cleaning and Disinfecting Your Facility’ to provide a general guidance regarding cleaning and disinfection at workplace to prevent the spread of SARS-CoV-2 virus that causes COVID-19 *(4)*. On April 22^nd^, 2020, the CDC published a memorandum based on its investigation of the Smithfield Foods COVID-19 outbreak to provide recommendations for improving health and safety controls at their production plant *(38)*. On April 26^th^, 2020, the CDC and the Occupational Safety and Health Administration (OSHA) jointly published the guidance documents ‘Meat and Poultry Processing Workers and Employers’ and ‘Protecting Seafood Processing Workers from COVID-19’ to assist the employers and workers specifically in the food processing industry in identifying risks associated with COVID-19 exposure and preventing virus spread at workplace *(7, 8)*. A number of other documents were also issued over a relatively short period of time. Therefore, there is a need for a collated list and review of those documents issued early in the response to the COVID-19 pandemic (i) to aid researchers and public health workers in interpreting potential timing of implementation of mitigation strategies and the attributed changes in the epidemiology of the disease in the food processing industry work environment, as well as, (ii) to guide the employers in the food processing industry in controlling COVID-19 exposure risks and maintaining facility operation. Thus, the objective of this study was to identify, review, and collate US government-published documents that contain resources or guidelines about COVID-19 mitigation relevant to the US food manufacturing and processing facilities.

## MATERIALS AND METHODS

### Document collection and search criteria

Rapid web-based review *(39)* was conducted between June 1^st^ and September 30^th^, 2021, to collect documents that contain resources or guidelines about COVID-19 mitigation published by government institutions, including the CDC, OSHA, and the Food and Drug Administration (FDA). Each of these three institutions has a dedicated COVID-19 webpage *(27, 45, 57)*, which we reviewed including all tabs under each page. A complementary snowballing approach was used to identify additional relevant documents from other US government agencies or professional associations where applicable. Collected documents were screened to identify those that (i) are relevant to the food manufacturing and processing industry workplace (i.e., food service or food retail industry was not within the scope of this review) and (ii) were accessible or published between the beginning of the pandemic and the end of the search period (September 30^th^, 2021). Frequently asked questions (FAQ) pages were not included in this review. Z.C. identified the documents, retrieved the publishing date for each document, read and subjected each document to content analysis (described in the next sections).

### Launch date data collection

The original publishing date of each document was retrieved through one of the following two methods. The first method involved going directly to the document webpage and retrieving the website source code *(51)*. In the source code, key words ‘first_published’ or ‘published_time’ represent the date when the website first went public and became accessible on the internet. In rare cases when the source code did not contain information regarding publishing date, the second method was used as it provides the publishing date of any website. This method involved using ‘Carbon Dating the Web’, a web application which estimates the webpage launch date by inputting the URL link of the target webpage *(48)*. Therefore, both methods together resulted in complete information about the date when each of the identified documents was first published.

### Content analysis

After the initial collecting and screening of documents, content analysis was conducted on all identified documents, and a codebook was developed in Microsoft Excel to record the mitigation strategies mentioned in each document. Specifically, the whole content of each document was extracted and coded based on the category of mitigation strategy. For that purpose, we adapted a list of categories of mitigation strategies from a recent needs assessment survey of COVID-19 perceptions in the food processing industry *(42)*, and added to the original list any new category that we came across in the documents. The list of considered categories and their detailed explanations are shown in the Supplemental Material (**Table S1**). After generating a list of categories of mitigations, the frequency of each category was also counted in terms of the number of reviewed documents mentioning a given category. Excel spreadsheets used to organize and analyze the reviewed documents can be accessed at https://github.com/IvanekLab/Control-of-COVID-19-in-the-US-food-industry.

## RESULTS AND DISCUSSION

This study aimed to survey documents published by the US government institutions regarding COVID-19 mitigation in the US food manufacturing and processing industry. A total of 36 publicly available documents were identified through a rapid review of the CDC, OSHA, FDA webpages, as well as snowballing to other relevant government agencies (the United States Department of Agriculture (USDA) and the Federal Emergency Management Agency (FEMA)) and to a single professional agency (the American Society of Heating, Refrigerating and Air-Conditioning Engineers (ASHRAE)). A timeline of when the 36 documents were published was created (**Figure 1**). As shown in the timeline, a cluster of documents was published in April 2020, following WHO’s declaration of COVID-19 as a global pandemic *(34)*, and coinciding with the outbreak at Smithfield Foods *(36)*. In December 2020, the second cluster of documents was published when a large spike in COVID-19 cases occurred in the USA during the winter *(52)*.

**Figure 1.**
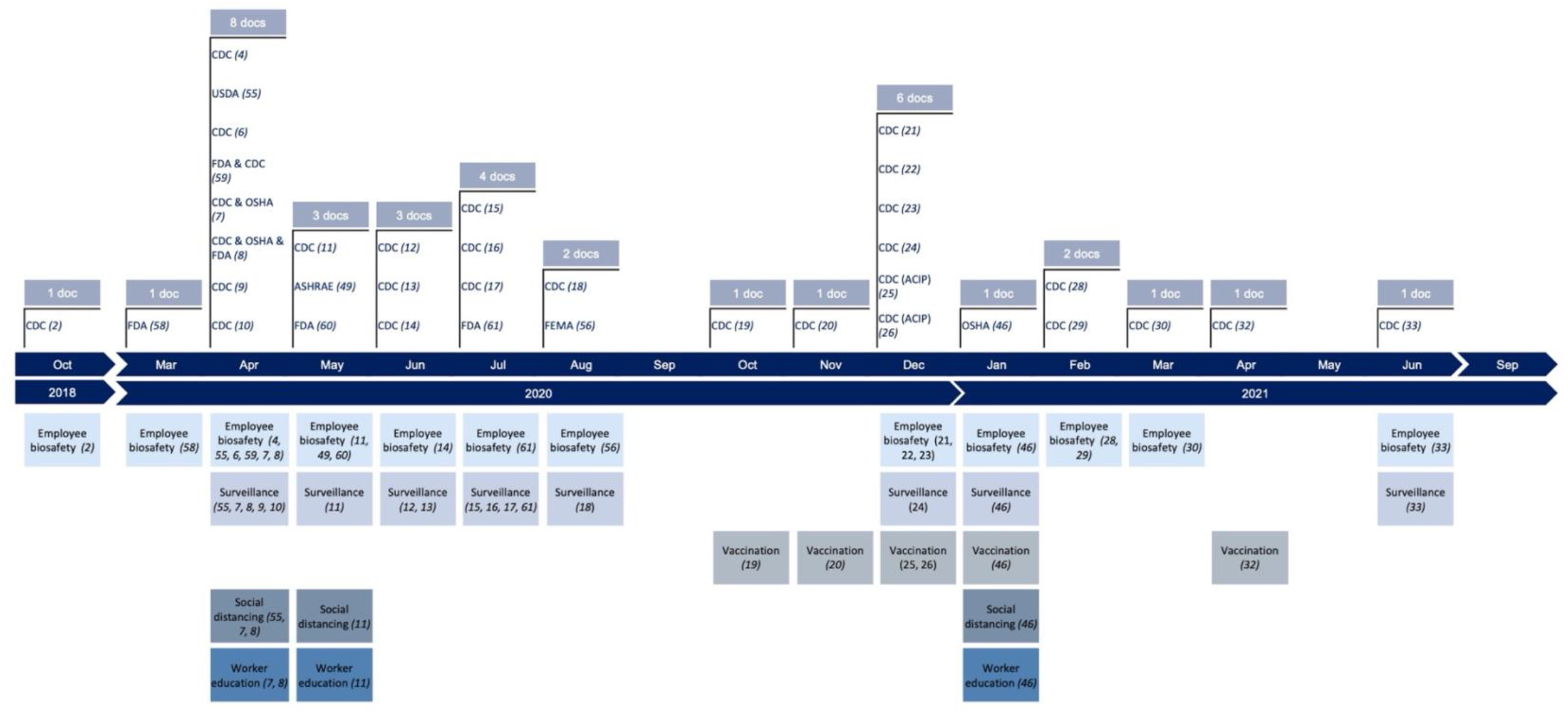
Timeline of the documents published by the United States (US) government and a single professional association about mitigation of Coronavirus Disease 2019 (COVID-19) relevant to the US food manufacturing and processing industry. Shown above the timeline axis are individual documents by their publishing entities (and reference number), while below the timeline axis are the themes of mitigation strategies extracted from all documents published in a given month (ordered by row from the most to least frequently appearing theme).

Among the 36 documents, a total of 5 themes and 19 categories of mitigation strategies were identified and extracted through content analysis (**Table 1**). Overall, the categories cover the broad themes of employee biosafety, surveillance, vaccination, social distancing (also known as “physical distancing”) *(5)*, and worker education. The first document describing the employee biosafety theme was published already in October 2018 (**Figure 1**), i.e., before the pandemic has started, but it was included in this review because of its relevance to the COVID-19 control. In April 2020, surveillance, social distancing, and worker education mitigation themes were first mentioned in the reviewed documents, while the vaccination theme was first mentioned in a document published in October 2020 (**Figure 1**). The frequency of each category of mitigation strategies appearing in the 36 documents is shown in **Table 2**. The frequency of appearance can be influenced by many factors, such as, but not limited to, changes in the scientific knowledge of the SARS-CoV-2 virus, ease and cost of implementation, or lack of compliance towards certain infection controls *(47, 64)*. Among the 19 categories, ‘Face mask, face shields, goggles,’ ‘Test for infection and isolation,’ and ‘Workplace cleaning and disinfection’ were each mentioned in 14, 11, and 8 out of the total 36 documents, ranking as the top three most frequently mentioned mitigation strategies. A high frequency of appearance may make it more difficult for the industry employees to keep track of the information about the mitigation strategies. Therefore, this study may help provide a more organized and convenient means for the readers to locate relevant documents of interest.

**Table 1.**
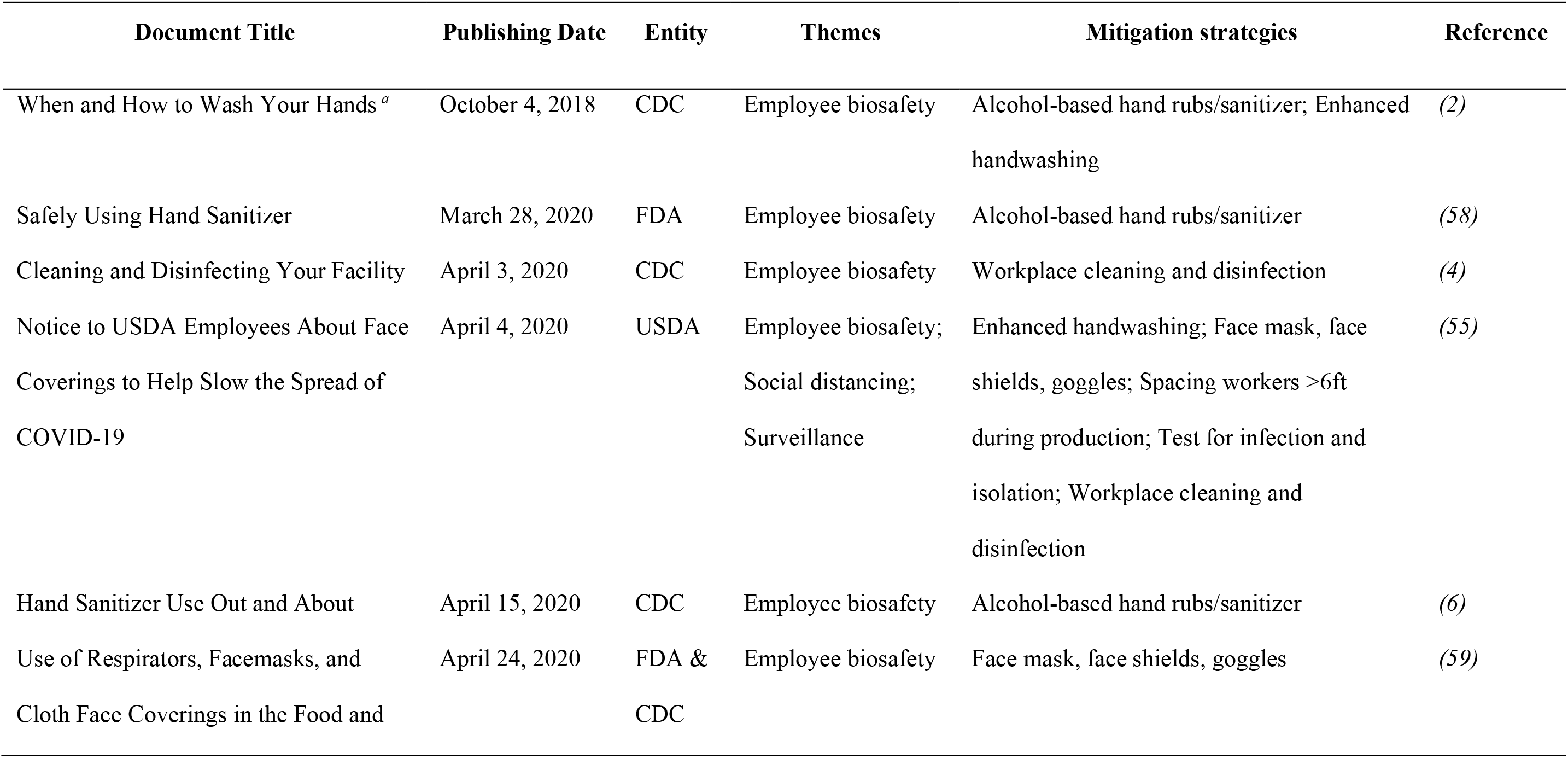
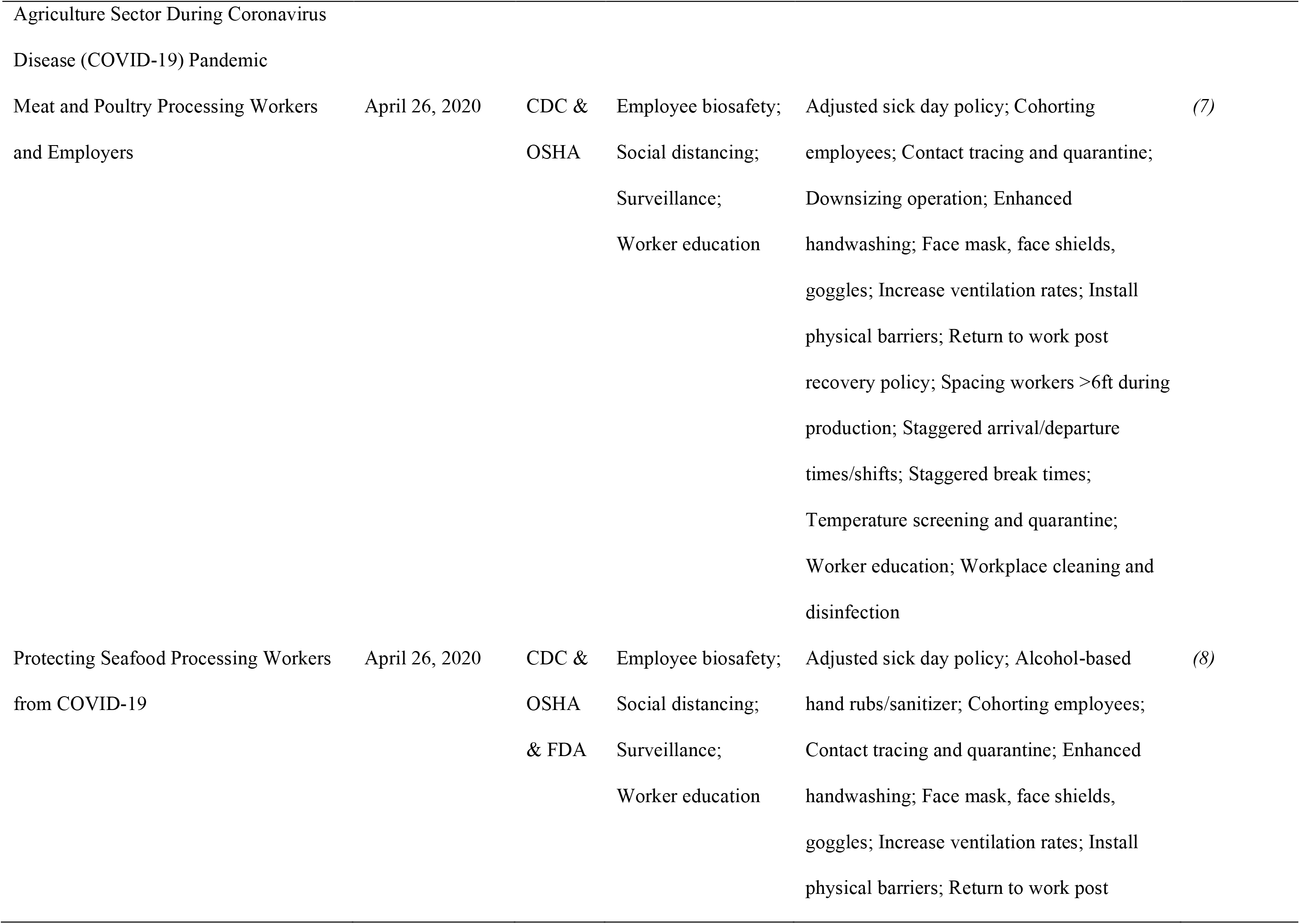
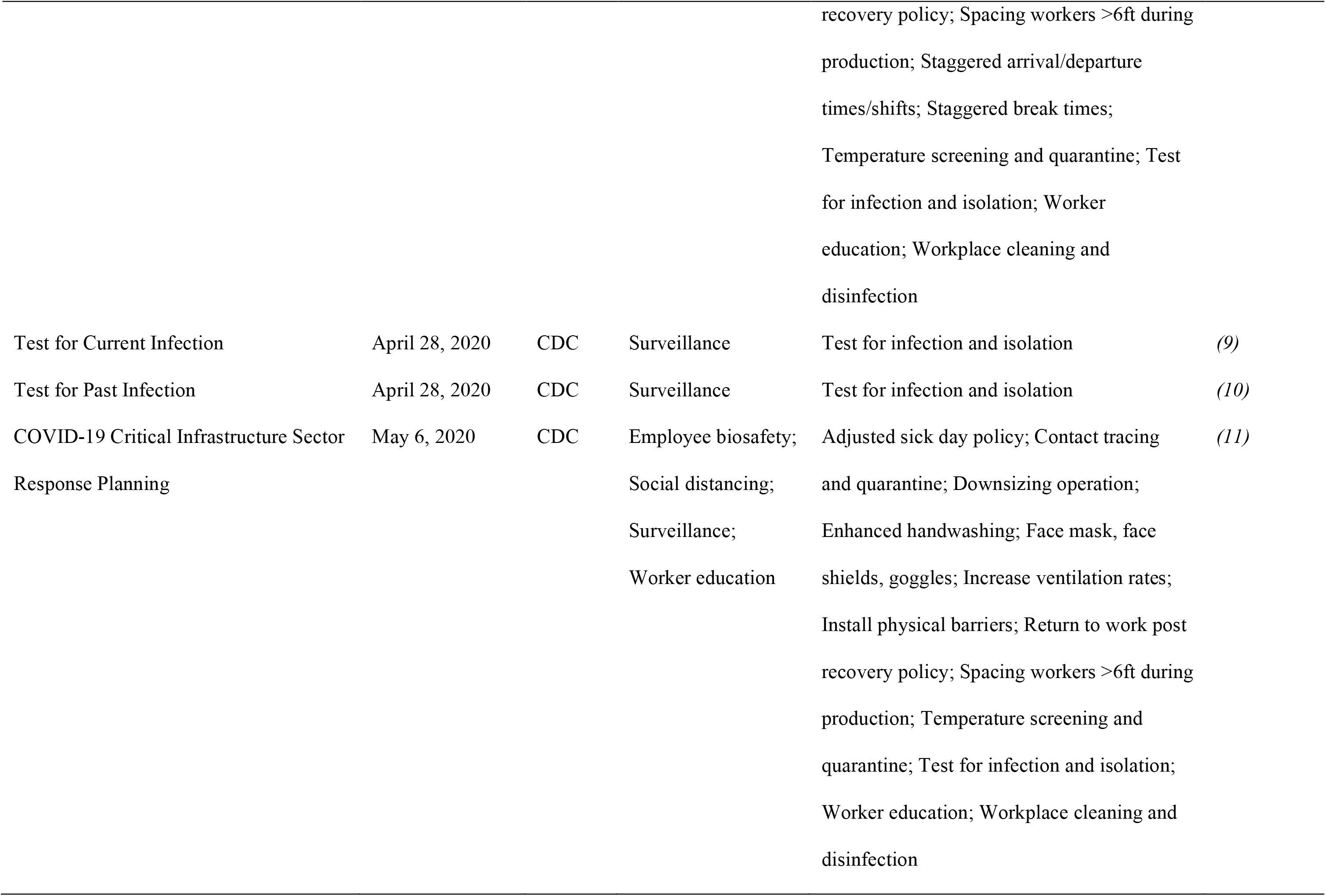
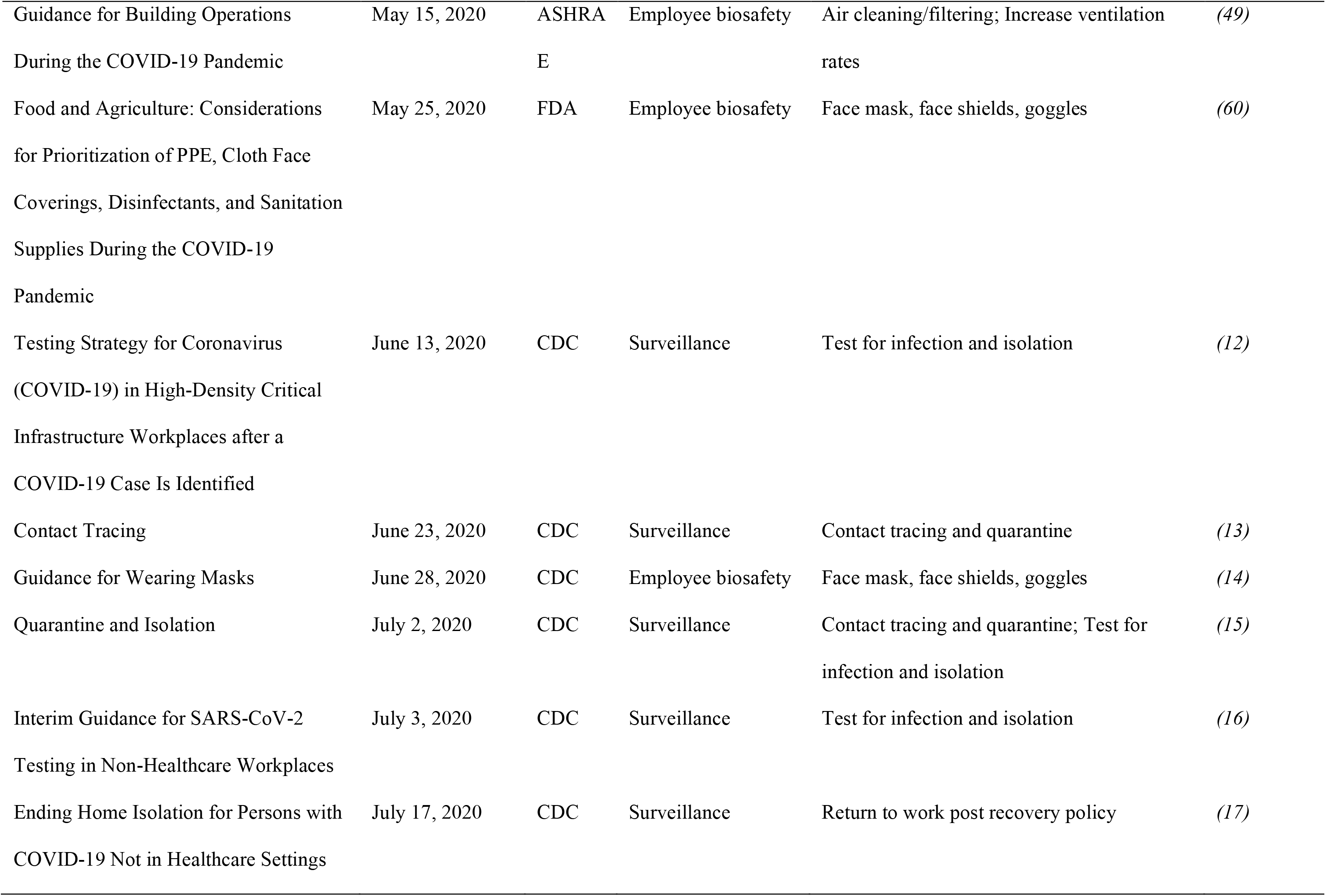
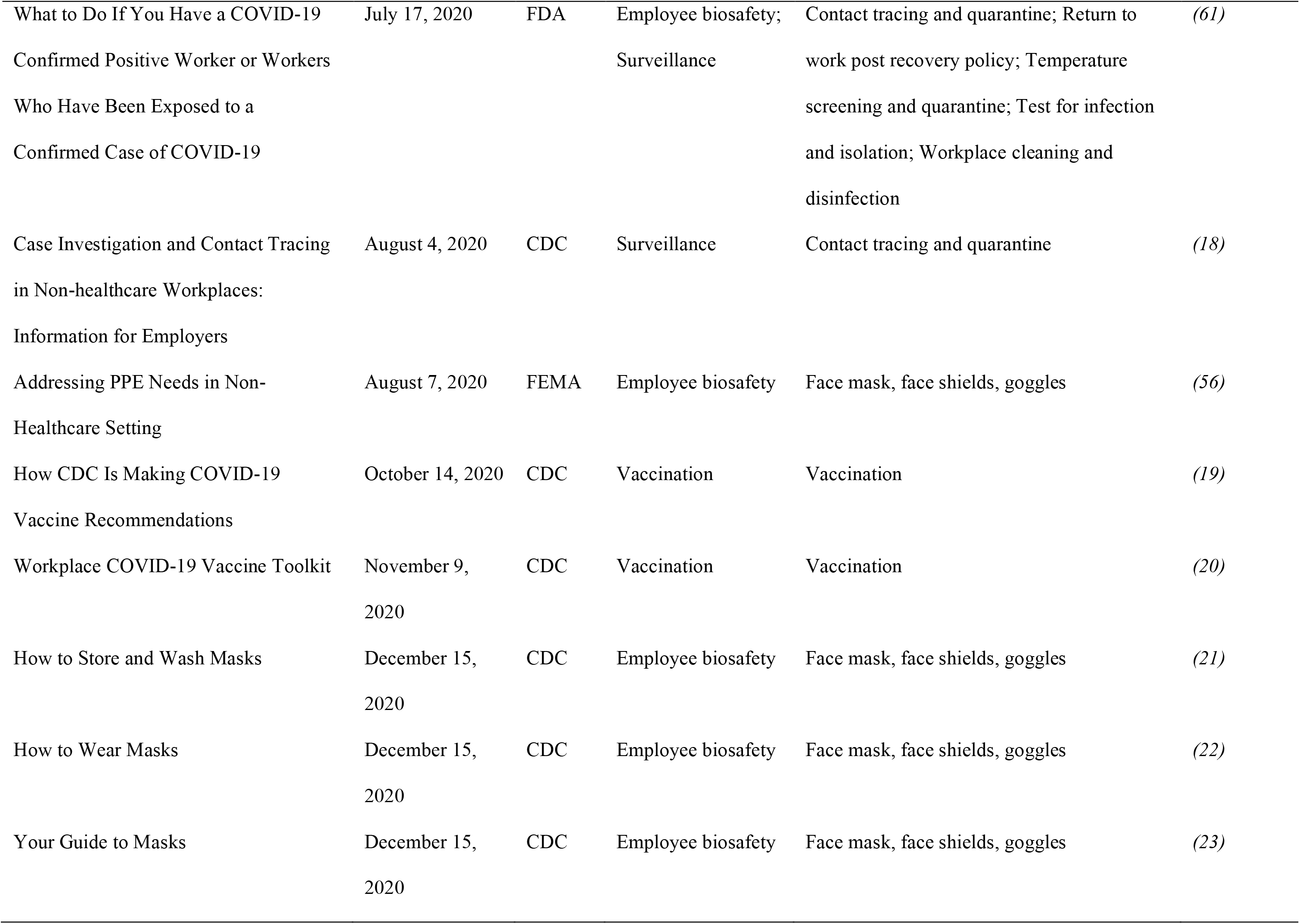
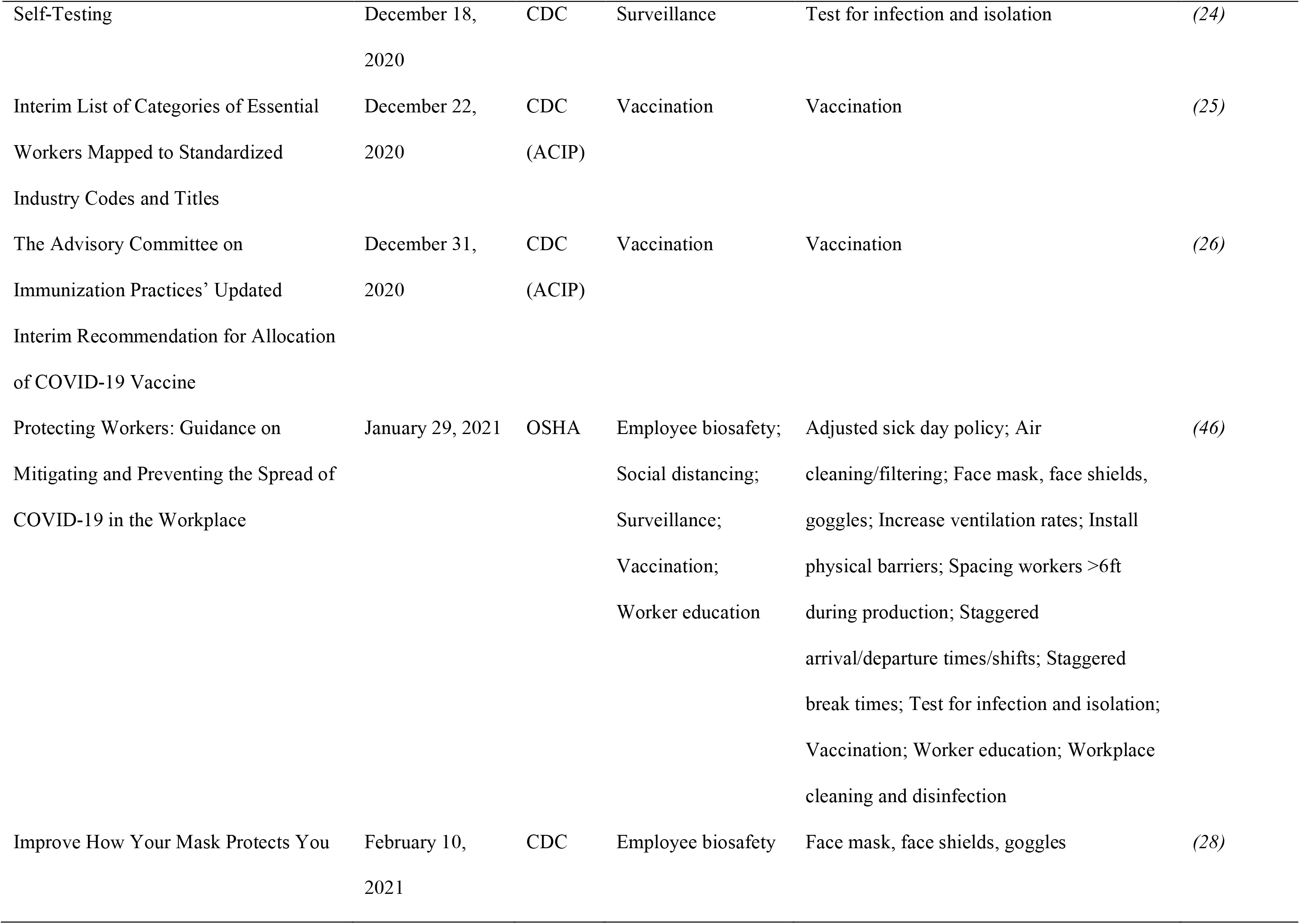
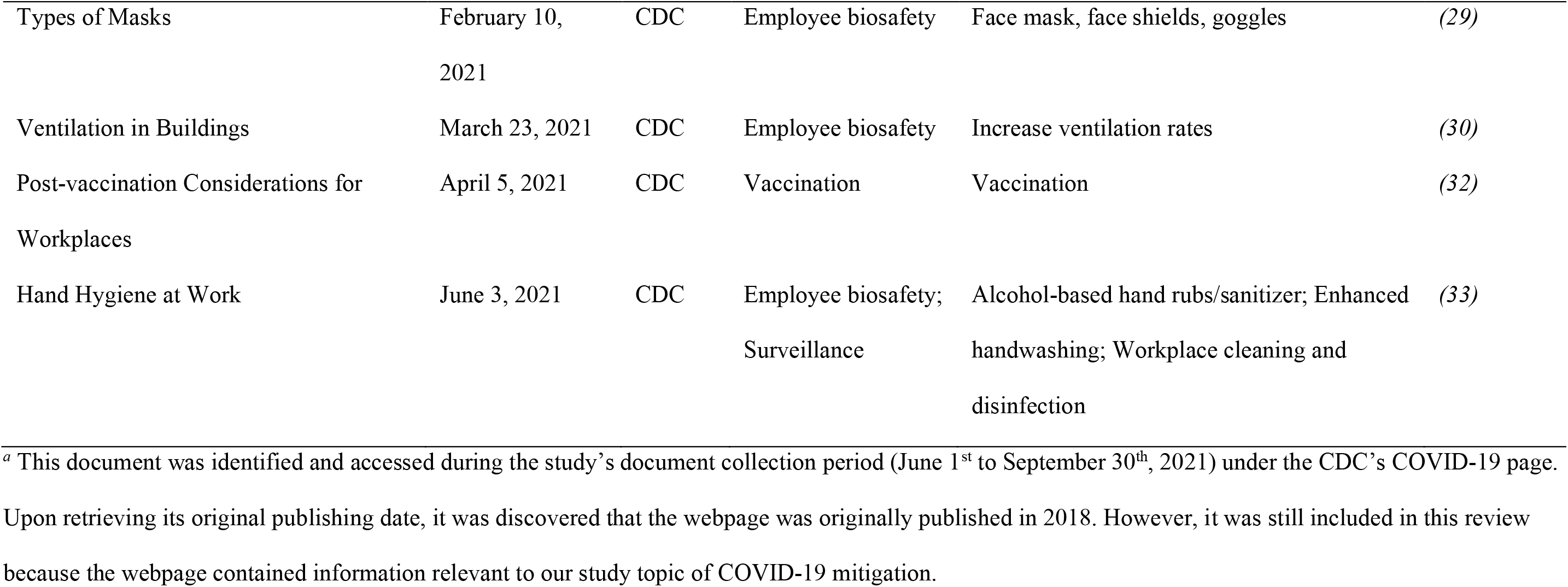
Identified documents published before September 30^th^, 2021, by the United States government and a single professional association on COVID-19 mitigation strategies relevant to the food manufacturing and processing industry

**Table 2.**
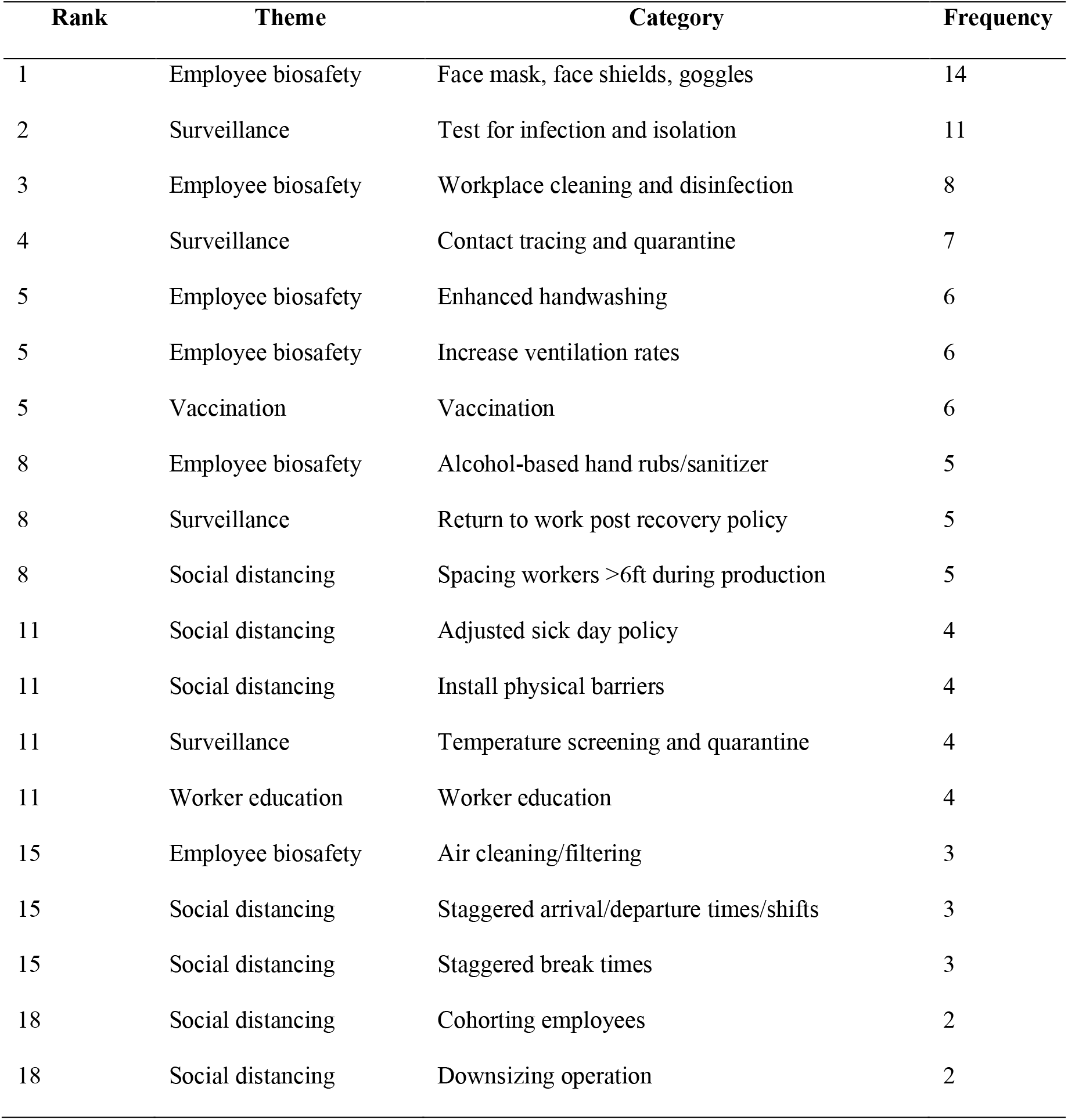
List of categories of mitigation strategies against COVID-19 in the United States food manufacturing and processing industry ranked by frequency of appearance in the reviewed documents

| Rank | Theme | Category | Frequency |
| --- | --- | --- | --- |
| 1 | Employee biosafety | Face mask, face shields, goggles | 14 |
| 2 | Surveillance | Test for infection and isolation | 11 |
| 3 | Employee biosafety | Workplace cleaning and disinfection | 8 |
| 4 | Surveillance | Contact tracing and quarantine | 7 |
| 5 | Employee biosafety | Enhanced handwashing | 6 |
| 5 | Employee biosafety | Increase ventilation rates | 6 |
| 5 | Vaccination | Vaccination | 6 |
| 8 | Employee biosafety | Alcohol-based hand rubs/sanitizer | 5 |
| 8 | Surveillance | Return to work post recovery policy | 5 |
| 8 | Social distancing | Spacing workers >6ft during production | 5 |
| 11 | Social distancing | Adjusted sick day policy | 4 |
| 11 | Social distancing | Install physical barriers | 4 |
| 11 | Surveillance | Temperature screening and quarantine | 4 |
| 11 | Worker education | Worker education | 4 |
| 15 | Employee biosafety | Air cleaning/filtering | 3 |
| 15 | Social distancing | Staggered arrival/departure times/shifts | 3 |
| 15 | Social distancing | Staggered break times | 3 |
| 18 | Social distancing | Cohorting employees | 2 |
| 18 | Social distancing | Downsizing operation | 2 |

### Multiple government institutions have issued documents relevant to the mitigation of COVID-19 among workers in the food processing industry

Since the start of COVID-19 outbreak in the USA in early March 2020, the CDC and other government institutions have acted quickly through responding and providing numerous documents containing resources or guidelines about COVID-19 mitigation to the general public as well as to different industries. Specifically for the food processing industry, the FDA published a dedicated site for the food and agriculture sectors on March 17^th^, 2020 *(57)*, and the CDC and OSHA jointly published the guidance documents ‘Meat and Poultry Processing Workers and Employers’ and ‘Protecting Seafood Processing Workers from COVID-19’ on April 26^th^, 2020 *(7, 8)*.

Overall, the CDC has been at the forefront of responding to the pandemic and served two major roles in COVID-19 mitigation. First, the CDC served as the main platform to provide COVID-19 related information, from disease surveillance and summary of scientific studies to mitigation strategies. This is expected considering the mandate of the CDC is to protect the nation from health threats *(3)*. Thus, the CDC provided detailed resources and guidelines in all aspects of disease control *(31)*. Besides general mitigation strategies, such as face masking, viral testing, vaccination, contact tracing, quarantine, and temperature screening (**Table 1**), the CDC has also suggested workplace specific strategies to help combat virus spread at food processing facilities, including increased ventilation, production line distancing, adjusted sick day policy, physical barriers, staggered break time and shifts, and worker cohorting *(4, 7, 8, 11, 28)*. The CDC’s second role was to serve as a reference point for other agencies to further develop more specialized guidelines. For example, the USDA and OSHA built their industry specific guidelines based upon the evidence-based documents that the CDC had developed *(46)*. These guidelines were critically important for the employers of the food processing industry as they relied on this information to implement and incorporate the mitigation strategies into their processing facilities’ daily routine to control COVID-19 infection at workplace.

As numerous documents have been released by different government agencies, our organized timeline of when each relevant document was published, and the themes and categories of mitigations covered, can be helpful in assessing the corresponding changes in the epidemiology of COVID-19 in the food processing industry work environment. In addition to the intended purpose to reduce the spread of infection, some of these short-term mitigation strategies, such as workplace disinfection and ventilation improvement, can have long-term benefits to the food processing industry as they could potentially further improve the hygiene standards at the facilities and promote establishing effective strategies that reduce contamination with foodborne pathogens during the food production process *(53)*. Additionally, having a plan developed for infectious diseases at workplace can help the food processing facilities in preparing for any similar pandemic in the future *(42)*.

### Overall goals were to ensure the safety of workers and maintain food supply

As the food and agriculture sectors were identified as critical infrastructure during the COVID-19 pandemic *(54)*, the two overarching goals of COVID-19 mitigation in the food processing industry workplace have been (i) to ensure the safety of the workers and (ii) to maintain the operation of food processing facilities as a critical infrastructure. As such, in one of the reviewed documents ‘COVID-19 Critical Infrastructure Sector Response Planning’, the CDC requires exposed workers in critical infrastructure to get tested but allows them to continue working as they remain asymptomatic and have not tested positive *(11)*. In face of a global pandemic, it is critically important to put the health of the food processing industry employees as the highest priority, while it is also necessary to maintain facility operation to satisfy public needs and to keep societal order. As the National Infrastructure Simulation and Analysis Center (NISAC) modeled in a previous study, a severe pandemic with 25% reduction in labor can reduce food production by almost 50%, causing food shortage *(41)*. In addition, particularly with the SARS-CoV-2 virus, a major practice to slow the spread of the infection has been social distancing at home, which would result in modified daily activities and potentially increased food demand *(1)*. Any imbalance between demand and supply can cause food price inflation, food shortage, and consequently food insecurity if no action is taken *(63)*. Therefore, as employers develop COVID-19 mitigation programs for their facilities, it is important to ensure the safety of the workers and maintain the operation of food processing facilities as it strengthens the food supply chain to avoid nation-wide, or even worse, world-wide food crisis.

### Study limitation

In line with the search period in this review, identified documents were limited to the period before October 2021. Consistent with the practice for rapid reviews *(35)* we focused on a specific segment of the food industry, and we adapted the search methodology to enable a rapid review process. Specifically, we thoroughly searched the CDC, OSHA, and FDA websites for food manufacturing and processing industry relevant COVID-19 information because we presumed that these three institutions are the most prominent government authorities relied on for guidance during the pandemic in the USA. Since the search methodology was not intended to provide an exhaustive review of all documents about COVID-19 mitigation ever published on the internet, relevant documents published by other government agencies and the corresponding additional mitigation strategies might have been missed. To address this, we expanded the search process with snowballing and identified a few additional documents in an effort to reduce the potential bias associated with the search methodology.

## CONCLUSION

This article provides a summary of the common types of COVID-19 mitigation strategies that have been recommended for implementation in the US food manufacturing and processing industry. Based on the documents collected in this review, it can be concluded that the government was able to respond within a couple of weeks to the pandemic by providing evidence-based resources and guidelines for the public. Specifically, ‘Face mask, face shields, goggles,’ ‘Test for infection and isolation,’ and ‘Workplace cleaning and disinfection’ were the three most frequently mentioned mitigation strategies of relevance to the food processing industry.

The summarized timeline of published documents can serve as a useful resource for public health researchers in studying implications of the implementation of different mitigation strategies in the food processing industry workplace, as well as for the food processing facilities to refer to as they plan for their COVID-19 management policy. During a pandemic, it is important that the food processing facilities protect their worker’s health and safety but maintain operations as much as possible to ensure an uninterrupted food supply chain and food security. It is a time that requires the facility management to make both engineering and policy changes to help adjust to the situation. Importantly, these changes can potentially bring positive long-term impacts to the food processing industry and prepare the food processing facilities for any similar situations in the future.

## Data Availability

All data referred to in the present study are contained in the manuscript; and the excel spreadsheets referred to in the present study are available online at

https://github.com/IvanekLab/Control-of-COVID-19-in-the-US-food-industry

## ACKOWLEDGEMENTS

This study was funded by the United States Department of Agriculture (USDA), National Institute of Food and Agriculture (NIFA) award #2020-68006-32875 to R.I. The funders had no role in study design, data collection and analysis, decision to publish, or preparation of the manuscript.

The authors thank Brianna Dumas, MPH, RD (Health Scientist, National Center for Chronic Disease Prevention and Health Promotion, CDC), CAPT John D Gibbins, DVM, MPH, Diplomate ACVPM (Team Lead, National Institute for Occupational Safety and Health, CDC), CDR Adam Kramer, ScD, MPH, RS (Environmental Health Officer, National Center for Environmental Health, CDC), and Arthur P Liang, MD, MPH (Special Advisor on Food Industry Relations, National Center for Emerging and Zoonotic Infectious Diseases, CDC) for providing helpful comments on the manuscript.

