## Supplemental Table 1 for "Rapid review of government issued documents relevant to mitigation of COVID-19 in the US food manufacturing and processing industry"

**SUPPLEMENTAL MATERIAL**

**Table S1.** Definitions of themes and categories of COVID-19 mitigation strategies considered in the content analysis

| | Theme | Category | Definition [a] |
|---|---|---|---|
| 1 | Employee biosafety | Air cleaning/filtering | Destroying or removing hazards like viral particles from air. |
| 2 | Employee biosafety | Alcohol-based hand rubs/sanitizer | Implementation of a set of instructions for employees about when and how to use alcohol-based hand rubs and sanitizer. |
| 3 | Employee biosafety | Enhanced handwashing | Implementation of a set of instructions for employees about when and how to wash hands that goes above and beyond instructions that were in place pre-COVID-19. |
| 4 | Employee biosafety | Face mask, face shields, goggles | Implementation of a set of instruction about how and when to use face masks, face shields and goggles. |
| 5 | Employee biosafety | Increase ventilation rates | Increase in the rate at which external air (fresh air) flows into the building. |
| 6 | Employee biosafety | Workplace cleaning and disinfection | Instructions on how to clean and disinfect the facilities as a daily routine or when there are workers diagnosed with COVID-19. |
| 7 | Social distancing | Adjusted sick day policy | Employee benefits include a paid sick leave granted when an employee is unable to work because the employee is quarantined or isolated due to COVID-19, because of a bona fide need to care for an individual subject to quarantine or isolation, or to care for a child (under 18 years of age) whose school or childcare provider is closed or unavailable for reasons related to COVID-19. |

| | | | |
|---|---|---|---|
| 8 | Social distancing | Cohorting employees | Establishing groups of employees based on their risk of infection in the company, where each cohort remains as separated from the other cohorts as possible. |
| 9 | Social distancing | Downsizing operation | Reduction of a facility's production capacity accompanied with a reduction in the number of employees. |
| 10 | Social distancing | Install physical barriers | Clear plastic partitions preventing employees from getting too close and preventing particles or droplets exhaled by one person from entering the breathing zone of another. |
| 11 | Social distancing | Spacing workers >6ft during production | Keeping a space at least 6 feet between employees. |
| 12 | Social distancing | Staggered arrival/departure times/shifts | Groups of employees have a set number of hours to work during the day, but they have different start and finish times. |
| 13 | Social distancing | Staggered break times | Groups of employees have different break times. |
| 14 | Surveillance | Contact tracing and quarantine | Contact tracing is a process to identify individuals who may have been exposed to a person with COVID-19. Quarantine is the practice of separating individuals who have had close contact with someone with COVID-19 to determine whether they develop symptoms or test positive for the disease. |
| 15 | Surveillance | Return to work post recovery policy | Any strategy implemented for employees returning to work following a COVID-19 infection based on symptoms or doctor's recommendation. |
| 16 | Surveillance | Temperature screening and quarantine | Screen for employees with temperature above 99.5°F (or other cut-off value) and keep identified employees away from workplace to determine whether they develop COVID-19 symptoms or test positive for the disease. |

| | | | |
|---|---|---|---|
| 17 | Surveillance | Test for infection and isolation | Test employees for COVID-19 infection (viral test); Isolation: keep away from workplace an employee who is sick with COVID-19 or tested positive for COVID-19 without symptoms. |
| 18 | Vaccination | Vaccination | Any recommendation regarding having the employees to be vaccinated against COVID-19. |
| 19 | Worker education | Worker education | Employers educate and train workers and supervisors about how they can reduce the spread of COVID-19 infection. |

[a] Definitions of mitigation categories were adapted from a study by Llanos-Soto, S., E. Bulut, S. I. Murphy, C. J. Henry, C. Zoellner, M. Wiedmann, D. Wetherington, A. Adalja, S. D. Alcaine, and R. Ivanek. 2021. Ongoing mitigation strategies and further needs of the United States food industry to control COVID-19 in the work environment. *medRxiv.* [Preprint.] Available at: https://doi.org/10.1101/2021.08.06.21261702.
